# Observational Study of the Efficiency of Treatments in Patients Hospitalized with Covid-19 in Madrid

**DOI:** 10.1101/2020.07.17.20155960

**Authors:** Nikolas Bernaola, Raquel Mena, Ander Bernaola, Cesar Carballo, Antonio Lara, Concha Bielza, Pedro Larrañaga

**Affiliations:** Universidad Politécnica de Madrid, Spain; Universidad Complutense de Madrid, Spain; Universidad Francisco de Vitoria, Madrid, Spain; Hospital Universitario Ramon y Cajal, Madrid, Spain; Hospital Universitario Sanitas La Zarzuela, Madrid, Spain

## Abstract

**Background:** Many different treatments were heavily administered to patients with COVID-19 during the peak of the pandemic in Madrid without robust evidence supporting them.

**Methods:** We examined the association between sixteen treatments in four groups (steroids, antivirals, antibiotics and immunomodulators) and intubation or death. Data were obtained from patients that were admitted to an HM hospital with suspicion of COVID-19 until 24/04/2020, excluding unconfirmed diagnosis, those who were admitted before the epidemic started in Madrid, had an outcome that was not discharge or death or died within 24 hours of presentation. We compared outcomes between treated and untreated patients using propensity-score caliper matching.

**Results:** Of 2,307 patients in the dataset, 679 were excluded. Of the remaining 1,645 patients, 263 (16%) died and 311 (18.9%) died or were intubated. Except for hydroxychloroquine and prednisone, patients that were treated with any of the medications were more likely to go through an outcome of death or intubation at baseline. After propensity matching we found an association between treatment with hydroxychloroquine and prednisone and better outcomes (hazard ratios with 95% CI of 0.83 ± 0.06 and 0.85 ± 0.03). Results were similar in multiple sensitivity analyses.

**Conclusions:** In this multicenter study of patients admitted with COVID-19 hydroxychloroquine and prednisone administration was found to be associated with improved outcomes. Other treatments were associated with no effect or worse outcomes. Randomized, controlled trials of these medications in patients with COVID-19 are needed to avoid heavy administration of treatments with no strong evidence to support them.

## Introduction

The coronavirus disease 2019 (COVID-19) pandemic, declared by the WHO Director General at the media briefing on March 11^th^ 2020, is caused by the named Severe Acute Respiratory Syndrome Coronavirus 2 (SARS-CoV-2)^1^. It started in Wuhan, China, but it later spread to the rest of the world, with currently over 8.7 million confirmed cases worldwide, most of them in Europe and North America^2^.

Spain, with over 246,000 confirmed cases and over 28,000 confirmed deaths as of June 22^nd^ 2020^2^, has one of the highest burdens of COVID-19 per inhabitant worldwide (over 524 cases per 100,000 inhabitants). The numerical data provided by the Spanish Health Ministry is becoming confusing, and the Financial Times regarded the figures as deeply compromised^3^. After a quick search in PubMed with the keywords “novel coronavirus”, “COVID-19” or “SARS-CoV2” and “Spain”, we have found no articles that describe the characteristics of infected patients.

Because it is a new epidemic, the specific mechanisms and pathophysiology remain elusive, and the risk factors for death have not been accurately defined (mainly age and comorbidities)^4^. No effective vaccine or antiviral treatment is currently available^5^. The analysis of data on the clinical characteristics, received treatment and outcomes of COVID-19 patients is of vital importance to reduce its mortality^6^. It will allow the identification of possible prognostic factors and provide preliminary data for the future development of management algorithms.

In this article we report on the clinical characteristics, previous history, received treatments and clinical outcome of 1,645 COVID-19 patients admitted to several hospitals in Madrid, Spain. The goal of this report is to identify those clinical profiles most likely to benefit from specific treatment and possible early clinical prognostic factors.

## Methods

### Study Design and participants

This is a multi-center, retrospective, observational study done with anonymized records provided by several HM hospitals in Madrid, Spain. These records included information of 2,307 patients admitted with a diagnosis of COVID POSITIVE or COVID PENDING, since the first officially recognized cases in Madrid (24/02/2020) to the last update to the database (24/04/2020). Patients are monitored up to discharge or death. The full description of the dataset is included in Appendix 1.

#### Exclusion criteria

Patients admitted before the first cases were declared in Madrid (24/02/2020), who had not yet reached an outcome (discharge or death) by 24/04/2020, transferred to a different hospital for admission, voluntarily discharged, with a “COVID PENDING” diagnosis or that were interned for less than 24 hours were excluded from the analysis. After exclusion there were 1,645 patients remaining. The exclusion process is explained in Figure 1.

**Figure 1:**
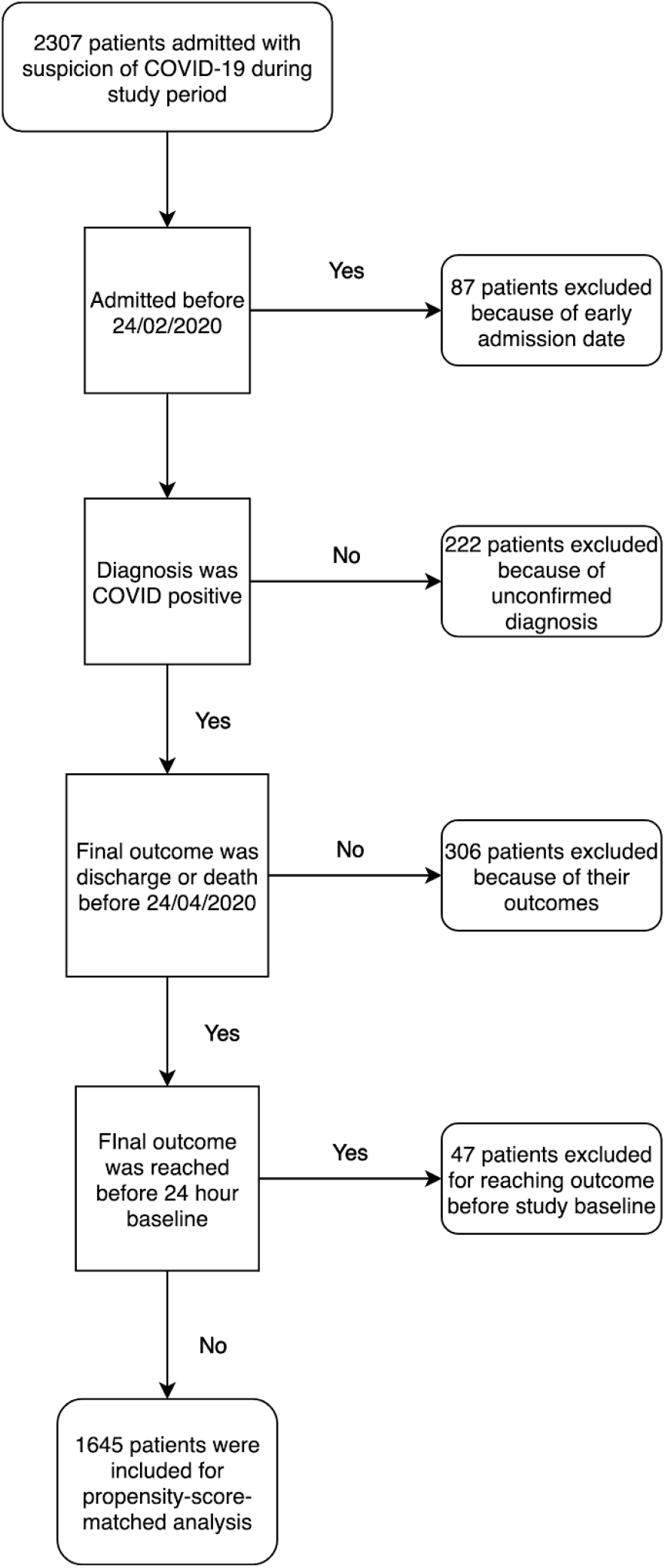
Flow diagram of the patients excluded from the intial sample

#### Clinical outcomes

In-hospital mortality was chosen as the primary outcome, defined as registered death of a patient before their discharge. As a secondary outcome, we chose a composite event that integrated both in-hospital mortality and intubation.

### Data sources

We obtained the data from the HM hospital network in Madrid, thanks to its project ‘COVID DATA SAVE LIVES’^7^. This anonymized clinical dataset comes from the HM hospitals HER system. It was openly released on April 25th on demand to any research groups that wanted to analyse it, provided they presented a project beforehand and said project was approved by the corresponding board of experts.

The data included patients’ age, sex, past diagnoses, smoking status, admission data, initial vital signs and complementary tests performed in the Emergency Room, vitals and tests performed throughout their hospital stay, treatments received (including previous medications continued and specific treatment for COVID-19), destination at discharge (or death) and diagnoses during their stay.

## Statistical analysis

### Variable selection

Since the dataset had an enormous number of variables for each patient, there were not always enough patients to effectively analyze each variable and obtain statistically significant associations while correcting for multiple comparisons and properly stratifying. Therefore, we chose to focus on only a few variables, as to avoid the statistical problems that come with high dimensional data. To do so, we consulted two independent experts on what the most important indicators for prognosis were. These variables are the following: age (divided into the following groups: <40, 40-59, 60-79 and ≥80), gender, past diagnoses (diabetes, hypertension, hypercholesterolemia, ischaemic events [recorded as continued use of anticoagulants and/or aspirin], chronic obstructive pulmonary disease [COPD] and cancer), toxic habits (current or past smoking), allergies (to penicillin and other medications), initial vital signs (temperature [measured in ^<u>°</u>^C], heart rate, oxygen saturation [measured through pulse oximetry] and blood pressure [systolic and diastolic, measured with an arm cuff]) and initial laboratory parameters (lactate dehydrogenase [LDH], D-dimer, ferritin, C-reactive protein [CRP], lactate, creatinine, procalcitonine, lymphocyte count and neutrophil count). For these features we took the first analysis they had after arriving at the hospital or, when possible, the average of the first two if taken within 24 hours so as to reduce the amount of missing data. Viral load and Interleukin-6 level were considered too but not enough patients were tested for them to be able to do the analysis. Viral load was not available in the data and only 30 patients had been tested for IL-6 level in their first two tests. All these variables and their distribution in our sample for patients that reached the primary outcome and those who did not are presented in Table 1. Table A2a shows the distribution stratified by the secondary outcome.

**Table 1:** Characteristics of the patients.

| Characteristics | Patients |  |  |
| --- | --- | --- | --- |
|  | Alive | Dead | Total |
| **Age - n (%)** |  |  |  |
| <40 | 107 (7.7) | 0 (0.0) | 107 (6.5) |
| 40-59 | 442 (32.0) | 6 (2.3) | 448 (27.2) |
| 60-79 | 638 (46.2) | 115 (43.7) | 753 (45.8) |
| $\geq 80$ | 195 (14.1) | 142 (54.0) | 337 (20.5) |
| **Female sex - n (%)** | 550 (39.8) | 83 (31.6) | 633 (38.5) |
| **Initial vital signs - median [Q1-Q3]** |  |  |  |
| Body temperature - °C | 36.7 (36.3-37.0) | 36.7 (36.3-37.0) | 36.7 (36.3-37.0) |
| Heart rate - beats/min | 90.0 (82.0-99.0) | 90.0 (80.0-96.5) | 90.0 (81.0-99.0) |
| Oxygen saturation - % | 94.0 (92.0-96.0) | 92.0 (84.0-94.0) | 93.0 (92.0-95.0) |
| Systolic blood pressure - mm Hg | 131.4 (125.0-136.8) | 131.4 (122.5-141.0) | 131.4 (125.0-138.0) |
| Diastolic blood pressure - mm Hg | 76.0 (72.0-80.5) | 75.3 (65.0-78.3) | 76.0 (71.0-80.0) |
| **Initial laboratory tests - median [Q1-Q3]** |  |  |  |
| Lactate dehydrogenase - U/L | 513 (411-610) | 660 (522-873) | 533 (419-646) |
| D-dimer - ng/mL | 972 (506-2002) | 1956 (1123-3350) | 1163 (546-2202) |
| Ferritin - ng/mL | 1148 (837-1477) | 1694 (1234-2389) | 1216 (868-1630) |
| C-reactive protein - mg/L | 50 (16-87) | 101 (59-189) | 58 (18-104) |
| Lactate - mmol/L | 1.7 (1.4-2.0) | 2.1 (1.8-2.9) | 1.7 (1.4-2.1) |
| Creatinine - mg/dL | 0.8 (0.7-1.0) | 1.1 (0.9-1.5) | 0.9 (0.7-1.0) |
| Procalcitonin -ng/mL | 0.0 (0.0-0.3) | 0.6 (0.2-1.3) | 0.1 (0.0-0.4) |
| Lymphocyte count per mm<sup>3</sup> | 1.2 (0.8-1.5) | 0.8 (0.5-1.2) | 1.1 (0.8-1.5) |
| Neutrophil count per mm<sup>3</sup> | 4.6 (3.2-6.2) | 6.8 (4.7-10.7) | 4.8 (3.4-6.8) |
| **Past diagnoses - n (%)** |  |  |  |
| Current smokers | 35 (2.5) | 6 (2.3) | 41 (2.5) |
| Current and past smokers | 181 (13.1) | 39 (14.8) | 220 (13.4) |
| Diabetes | 154 (11.1) | 52 (19.8) | 206 (12.5) |
| Hypertension | 393 (28.4) | 102 (38.8) | 495 (30.1) |
| Hypercholesterolemia | 266 (19.2) | 75 (28.5) | 341 (20.7) |
| Continued use of anticoagulants | 63 (4.6) | 32 (12.2) | 95 (5.8) |
| Continued use of aspirin | 101 (7.3) | 48 (18.3) | 149 (9.1) |
| Chronic obstructive pulmonary disease | 40 (2.9) | 20 (7.6) | 60 (3.6) |
| Cancer | 57 (4.1) | 29 (11.0) | 86 (5.2) |
| Penicillin allergy | 28 (2.0) | 9 (3.4) | 37 (2.2) |
| Other allergies to medication | 36 (2.6) | 5 (1.9) | 41 (2.5) |
| **Number of patients - n (%)** | **1382 (84.0)** | **263 (16.0)** | **1645 (100.0)** |

To analyze the differences in the distributions between patients that reached one of the outcomes and those who did not we conducted Mann-Whitney U tests for the continuous variables and Chi-square tests for the categorical ones. The *p*-values for the differences in the distributions both for the totality of the patients and stratified by age are shown in Table A3. After Bonferroni corrections to adjust for multiple comparisons we required *P* < .001 for statistical significance.

### Analysis

The main objective of our analysis was to examine the association between treatment and death rate for 16 drugs of interest. Two independent experts were prompted to choose which medication they considered critical from the 447 ATC7 identifiers in the original dataset, previously screened to include only the ones administered to at least 50 patients. The chosen drugs were of one of the following types: steroids, antivirals, antibiotics and immunomodulators. Both experts marked the same 16 drugs as critical. The full list of medication is reported in Table 2 along with the results. Appendix 9 is a short literature review of the chosen treatments with a summary of previous results and reasons why these treatments were considered useful for patients diagnosed with COVID-19.

An initial crude univariate analysis was done in which the treated and untreated groups for each medication were compared according to their outcomes. Due to the non-randomized nature of assignment of treatment, we then used a propensity-score based method^8^ to reduce the effects of confounding.

The primary analysis used was propensity-score caliper matching^9^. We first trained a logistic regression model using the variables in Table 1 to predict whether the patient took the medication or not, then we used the predicted probabilities for each patient as propensity scores for the respective medication. Table A4 shows the AUROC and AUPRC for these models and Appendix 5 shows the propensity-score distributions for all 16 treatments. Then, we used a caliper of 0.2 of the standard deviation in the propensity score of the population as recommended in the literature^10,11^ to match patients one-to-many from the smallest group (treated or untreated, depending on the medication) with patients from the other group with replacement, that is, we allowed the same patient from the bigger group to be matched to more than one patient of the smaller group. Patients that did not have any matches were discarded, although this was almost never the case and the worst case only excluded 6 patients, median exclusion rate at this caliper size was 1.1%. (See Table A7 for sensitivity analysis on caliper size and exclusion rates. Caliper size did not have a big effect in the results as long as the population of unmatched patients was not too big). Using these sets of matched patients, we calculated the weighted difference in means and effect sizes which we report as hazard ratios and Cohen’s d in Table 2 with 95% confidence intervals. For Cohen’s d we apply the usual interpretation^12^ where an effect size of less than 0.2 is considered negligible, 0.2 to 0.5 is small, 0.5 to 0.8 is medium and more than 0.8 is large. All the analyses were done using Python v3.6.1 with the sklearn, numpy, pandas and scipy libraries.

## Results

### Characteristics of the patients

Of the 2,307 patients in the original dataset (last updated 24/04/2020), only 2098 had been resolved. Following the exclusion criteria described above, 1645/ rema//ined for analysis. Their characteristics are shown in Table 1. Over the two-month /period of the study, 263 patients (16%) died and 1382 were discharged. For the initial laboratory tests and initial vital signs, missing data was imputed using multiple iterated imputation^13^. Appendix 2 shows the results of the statistical analysis considering a composite event (intubation or death) instead of only death. Appendix 3 shows the results of hypothesis tests for the association between these characteristics and both chosen outcomes (death and death or intubation), firstly considering the whole sample and afterwards stratifying by the two eldest age groups (60-79 years and ≥80years) since the other two did not have enough patients that underwent primary or secondary outcomes.

Among the initial vital signs, the main difference between the patients that died and those who did not, was the initial oxygen saturation. When analysing the difference in the distributions, it was the only parameter with *P*<.001 in all groups. Diastolic blood pressure also showed a significant effect, but this disappeared when separately considering the eldest age groups.

As for the initial laboratory tests, the analysed variables were chosen by experts according to their relevance for assessing risk, and their distributions are clearly distinct between the groups. After performing statistical hypothesis tests, all of these laboratory parameters showed statistically significant association (*P*<.001) with both outcomes chosen for our study. Only for D-dimer and when stratifying our sample according to age, did this significance disappear in the eldest group (≥80 years) (*P*=.02 and *P*=.04 for the primary and secondary outcomes respectively).

Finally, our data confirmed some previous clinical history as risk factors for a higher mortality rate, as they have likewise been proven to be in numerous other diseases, also with statistical significance in most of them although this effect was mostly explained by age, since it disappeared when stratifying. Noticeably, smoking, hypertension and medication allergies did not show a significant association with any of the chosen outcomes even in the whole population. In general, patients with worse clinical outcome had a bigger disease burden at baseline and, therefore, a smaller physiologic reserve.

#### Effect of treatments in death rate

Table 2 shows the crude and propensity-matched hazard ratios for 16 medications, with their ATC7 identifier, given to COVID-19 patients, along with the estimated effect sizes. To estimate the effects we use hazard ratios with 95% confidence intervals and to show the size of the effect relative to the total variation between patients, we use Cohen’s d^14^ also with 95% confidence intervals.

In the initial crude analysis, all medications except for prednisone, ritonavir and lopinavir, azithromycin and hydroxychloroquine, were associated with higher mortality. Of these four, associated with a reduction in the mortality rate, only hydroxychloroquine had a medium positive effect (d= −0.58 ± 0.17).

But treatment administration is not random, since patients that receive specific treatment for any disease are usually those with a more severe clinical presentation. Therefore, we performed a second analysis applying propensity-score matching methods. In general, effect sizes for all medications were attenuated, except for prednisone. Its influence on outcome increased from a non-significant to a small positive value (hazard ratio 0.85 ± 0.06 and d= −0.42 ± 0.18). Hydroxychloroquine maintained a similarly positive effect despite its attenuation due to the propensity-score matched analysis, with hazard ratio 0.84 ± 0.08 and d = −0.44 ± 0.17.

Regarding the remaining medications received by the patients in our study, after propensity-matching, nine of them (methylprednisolone, ritonavir and lopinavir, oseltamivir, amoxicillin, azithromycin, ceftriaxone, levofloxacin, tocilizumab and interferon beta 1-b) had no significant effect on mortality (d<0.2). Two of them (dexamethasone and piperacillin) had a small negative effect (0.2<d<0.5), slightly increasing mortality. The remaining three drugs (hydrocortisone, linezolid and meropenem) had medium or large negative effects (d>0.5).

When comparing these results with the analysis according to composite critical event, as appears in Table A2b, the difference is minimal, with the only change being that amoxicillin goes to d=0.21± 0.23, a small negative effect. As an added step to check the robustness of the results we carried out an analysis with nearest-neighbour matching, where each patient in the smaller group (treated or untreated) was matched with the patient with the closest characteristics but different treatment value with no replacement. Table A6 shows the results. In general, this increased the confidence intervals of the effect sizes since the samples were usually smaller. The results were similar and mostly within error or with overlapping confidence intervals of the results in Table 2, with the biggest differences found for methylprednisolone and tocilizumab which both showed a small negative effect not within error of their results in Table 2.

**Table 2:** Hazard ratio with 95% confidence intervals and Cohen’s d for various treatments before and after propensity-score matching, for their effects on mortality rate. Negative d or hazard ratio less than 1 implies patients treated with that medication died less than those that did not. The opposite is true for positive d or >1 hazard ratio. Medications highlighted in red have at least a small positive d (>0.2) after propensity-score matching. Similarly, medications highlighted in green have d < −0.2.

| Treatment |  | Crude analysis |  | Propensity-score matching |  |
| --- | --- | --- | --- | --- | --- |
| Name | Patients treated (%) | Hazard ratios | Effect size (Cohen's d) | Hazard ratios | Effect size (Cohen's d) |
| **Steroids** |  |  |  |  |  |
| Methylprednisolone | 601 (36.5) | 1.11 ± 0.04 | 0.29 ± 0.10 | 1.01 ± 0.04 | 0.03 ± 0.10 |
| Dexamethasone | 221 (13.4) | 1.26 ± 0.07 | 0.72 ± 0.14 | 1.12 ± 0.07 | 0.34 ± 0.14 |
| Hydrocortisone | 63 (3.8) | 1.44 ± 0.12 | 1.21 ± 0.26 | 1.30 ± 0.14 | 0.82 ± 0.25 |
| Prednisone | 132 (8.0) | 0.94 ± 0.06 | -0.16 ± 0.18 | 0.85 ± 0.06 | -0.42 ± 0.18 |
| **Antivirals** |  |  |  |  |  |
| Ritonavir and Lopinavir | 1048 (63.7) | 0.93 ± 0.04 | -0.18 ± 0.10 | 0.97 ± 0.03 | -0.07 ± 0.10 |
| Oseltamivir | 124 (7.5) | 1.05 ± 0.07 | 0.12 ± 0.18 | 1.02 ± 0.07 | 0.05 ± 0.18 |
| **Antibiotics** |  |  |  |  |  |
| Amoxicillin | 77 (4.7) | 1.13 ± 0.10 | 0.36 ± 0.23 | 1.07 ± 0.10 | 0.19 ± 0.23 |
| Piperacillin | 85 (5.2) | 1.30 ± 0.11 | 0.83 ± 0.22 | 1.16 ± 0.12 | 0.42 ± 0.22 |
| Linezolid | 86 (5.2) | 1.51 ± 0.10 | 1.38 ± 0.22 | 1.30 ± 0.12 | 0.82 ± 0.22 |
| Azithromycin | 950 (57.8) | 0.95 ± 0.04 | -0.15 ± 0.10 | 0.93 ± 0.03 | -0.18 ± 0.10 |
| Ceftriaxone | 1024 (62.2) | 1.07 ± 0.03 | 0.19 ± 0.10 | 1.00 ± 0.03 | 0.01 ± 0.10 |
| Meropenem | 139 (8.4) | 1.46 ± 0.08 | 1.26 ± 0.18 | 1.28 ± 0.09 | 0.78 ± 0.18 |
| Levofloxacin | 405 (24.6) | 1.06 ± 0.04 | 0.17 ± 0.11 | 1.02 ± 0.04 | 0.05 ± 0.11 |
| **Immunomodulators** |  |  |  |  |  |
| Hydroxychloroquine | 1498 (91.1) | 0.79 ± 0.08 | -0.58 ± 0.17 | 0.84 ± 0.08 | -0.44 ± 0.17 |
| Tocilizumab | 309 (18.8) | 1.12 ± 0.05 | 0.32 ± 0.12 | 0.99 ± 0.05 | -0.01 ± 0.12 |
| Interferon beta 1-b | 96 (5.8) | 1.12 ± 0.09 | 0.32 ± 0.21 | 1.01 ± 0.10 | 0.03 ± 0.21 |

## Discussion

In this analysis of 1645 patients hospitalized with COVID-19 we find that the risk of death was significantly lower for patients treated with azithromycin, prednisone and, especially, hydroxychloroquine. The confidence intervals for these treatments overlap with those found in previous observational studies^15^. When comparing our results with a small review of the current literature about effectiveness of treatments for COVID-19, we found evidence in favour of hydroxychloroquine, with good virological and clinical outcomes in some studies, both alone and in combination with azithromycin, although these results were not uniform in all studies^16^. There were no studies found for standalone use of antibiotics, and regarding antivirals, we found no evidence of benefits that would recommend their use^17^. Finally, there is evidence in favour of tocilizumab^18^ and interferon beta 1b^19^, but mostly for patients in critical state. The evidence regarding corticosteroids is controversial^20^, with no significant proof of positive or negative effect, although they appear to improve outcome in critical cases^21^.

However, the effects of hydroxychloroquine and prednisone are medium after matching, and persist even when considering intubated patients (Appendix 2). We think this is at least small evidence in favor of hydroxychloroquine and prednisone having a positive effect in the treatment of COVID-19.

We find that risk factors in our data are in agreement with what is already common knowledge about COVID-19: age, comorbidities (mainly cardiovascular risk factors: Diabetes, hypertension, ischaemic coronary disease [measured through use of anticoagulants or aspirin]; as well as chronic obstructive pulmonary disease). We also find that smokers are less prevalent in our sample than in the Spanish population at large, indicating some measure of resistance to infection. However, once admitted in the hospital, their outcomes are not significantly different in any way we can measure from the overall population.

Also in accordance with expert opinion, no aspects of the initial clinical presentation except for O2Sat were found to be associated with a higher risk for a bad prognosis. However, laboratory parameters associated with systemic inflammatory response (lactate, LDH, D-dimer, ferritin and neutrophil count) did prove to have a relationship with higher mortality and intubation rates. A lower initial lymphocyte count and worse kidney function (measured through plasmatic creatinine) was also found in deceased and intubated patients. These parameters might be useful for an initial estimation of a patient’s prognosis, but these results alone do not give enough information to alter clinical practice.

## Data Availability

Data is available on request from the HM hospital network COVID data save lives by sending a project proposal and being approved.

https://www.hmhospitales.com/coronavirus/covid-data-save-lives

### Appendix 1: HM dataset description

*The following text has been copied from a document released by the HM hospital network along with its database explaining its project and the structure of the dataset*.

The information is organized in tables according to their content, all of them linked by a unique admission identifier. This identifier is the de-anonymization key, explicitly created for this purpose, and has nothing to do with the actual identifier of each admission.

1. The main table includes data on the admission and the patient (age and sex), data on the previous emergency if there has been one (2,226 records), data on their stay in the ICU if there has been one and records of the first and last set of emergency constants.
2. The medication table shows all the medication administered to each patient during admission (more than 60,000 records), with the dates corresponding to the first and last administration of each drug, identified by their brand name and classification in the ATC5/ATC7.
3. In the table of vital signs, there are all the basic records of constants (54,000 records so far) collected during admission with their date and time of registration.
4. The laboratory table contains the results of the determinations (398,884 records) of all the requests made to each patient during admission and in the previous emergency, if any.
5. And finally, the ICD10 coding tables show the records of diagnostic and procedural information coded according to the international ICD10 classification in its latest distributed version (does not include COVID), for the patients referred, both for episodes of hospital admission (more than 1,600) and for the emergency (more than 1,900) prior to those episodes, if any.

### Appendix 2: Results by critical composite event

#### Characteristics of patients

**Table A2a:** Characteristics of the patients divided depending on whether they underwent composite critical event (death or intubation) or not.

| Characteristics | Patients |  |  |
| --- | --- | --- | --- |
|  | No event | Critical event | Total |
| **Age - n (%)** |  |  |  |
| <40 | 101 (7.6) | 6 (1.9) | 107 (6.5) |
| 40-59 | 423 (31.7) | 25 (8.0) | 448 (27.2) |
| 60-79 | 618 (46.3) | 135 (43.4) | 753 (45.8) |
| ≥80 | 192 (14.4) | 145 (46.6) | 337 (20.5) |
| **Female sex - n (%)** | 533 (40.0) | 100 (32.2) | 633 (38.5) |
| **Initial vital signs - median (IQR)** |  |  |  |
| Body temperature - °C | 36.7 (36.3-37.0) | 36.7 (36.3-37.2) | 36.7 (36.3-37.0) |
| Heart rate - beats/min | 90.0 (82.0-99.0) | 90.0 (81.0-99.0) | 90.0 (81.0-99.0) |
| Oxygen saturation - % | 94.0 (92.4-96.0) | 92.0 (85.0-94.0) | 93.0 (92.0-95.0) |
| Systolic blood pressure - mm Hg | 131.4 (125.0-136.0) | 131.4 (123.0-140.0) | 131.4 (125.0-138.0) |
| Diastolic blood pressure - mm Hg | 76.0 (72.0-80.2) | 75.8 (66.5-79.0) | 76.0 (71.0-80.0) |
| **Initial laboratory tests - median (IQR)** |  |  |  |
| Lactate dehydrogenase - U/L | 511.2 (408.5-607.0) | 647.9 (518.9-865.7) | 533.0 (418.9-646.3) |
| D-dimer - ng/mL | 943.0 (501.5-1986.5) | 1887.0 (1003.5-3180.2) | 1162.9 (546.0-2201.5) |
| Ferritin - ng/mL | 1144.9 (833.7-1471.2) | 1648.4 (1185.1-2324.5) | 1216.2 (868.0-1630.2) |
| C-reactive protein - mg/L | 49.3 (15.6-87.4) | 92.3 (53.0-179.4) | 57.5 (18.4-103.8) |
| Lactate - mmol/L | 1.7 (1.4-2.0) | 2.1 (1.7-2.8) | 1.7 (1.4-2.1) |
| Creatinine - mg/dL | 0.8 (0.7-1.0) | 1.0 (0.8-1.4) | 0.9 (0.7-1.0) |
| Procalcitonin -ng/mL | 0.0 (0.0-0.3) | 0.6 (0.2-1.3) | 0.1 (0.0-0.4) |
| Lymphocyte count per mm<sup>3</sup> | 1.2 (0.8-1.5) | 0.8 (0.5-1.2) | 1.1 (0.8-1.5) |
| Neutrophil count per mm<sup>3</sup> | 4.5 (3.2-6.1) | 6.6 (4.5-10.2) | 4.8 (3.4-6.8) |
| **Past diagnoses - n (%)** |  |  |  |
| Current smokers | 34 (2.5) | 7 (2.3) | 41 (2.5) |
| Current and past smokers | 177 (13.3) | 43 (13.8) | 220 (13.4) |
| Diabetes | 148 (11.1) | 58 (18.6) | 206 (12.5) |
| Hypertension | 385 (28.9) | 110 (35.4) | 495 (30.1) |
| Hypercholesterolemia | 258 (19.3) | 83 (26.7) | 341 (20.7) |
| Continued use of anticoagulants | 62 (4.6) | 33 (10.6) | 95 (5.8) |
| Continued use of aspirin | 98 (7.3) | 51 (16.4) | 149 (9.1) |
| Chronic obstructive pulmonary disease | 38 (2.8) | 22 (7.1) | 60 (3.6) |
| Cancer | 53 (4.0) | 33 (10.6) | 86 (5.2) |
| Penicillin allergy | 27 (2.0) | 10 (3.2) | 37 (2.2) |
| Other allergies to medication | 35 (2.6) | 6 (1.9) | 41 (2.5) |
| **Number of patients - n (%)** | 1334 (81.1) | 311 (18.9) | 1645 (100.0) |

#### Treatment hazard ratios for critical event patients

**Table A2b:** Hazard ratio with 95% confidence intervals and Cohen’s d for various treatments before and after propensity-score matching, for their effect on composite critical event (intubation or death). Negative d or hazard ratio less than 1 implies patients treated with that medication died less than those that did not. The opposite is true for positive d or >1 hazard ratio. Medications highlighted in red have at least a small positive d (>0.2) after propensity-score matching. Similarly, medications highlighted in green have d < −0.2.

| Treatment |  | Crude analysis |  | Propensity matching |  |
| --- | --- | --- | --- | --- | --- |
| Name | Patients treated (%) | Hazard ratios | Effect size (Cohen's d) | Hazard ratios | Effect size (Cohen's d) |
| **Steroids** |  |  |  |  |  |
| Methylprednisolone | 601 (36.5) | 1.12 ± 0.04 | 0.32 ± 0.10 | 1.00 ± 0.04 | -0.00 ± 0.10 |
| Dexamethasone | 221 (13.4) | 1.34 ± 0.07 | 0.86 ± 0.14 | 1.11 ± 0.08 | 0.28 ± 0.14 |
| Hydrocortisone | 63 (3.8) | 1.51 ± 0.12 | 1.31 ± 0.26 | 1.17 ± 0.15 | 0.44 ± 0.25 |
| Prednisone | 132 (8.0) | 1.01 ± 0.07 | 0.02 ± 0.18 | 0.88 ± 0.08 | -0.31 ± 0.18 |
| **Antivirals** |  |  |  |  |  |
| Ritonavir and Lopinavir | 1048 (63.7) | 0.97 ± 0.04 | -0.08 ± 0.10 | 0.98 ± 0.03 | -0.06 ± 0.10 |
| Oseltamivir | 124 (7.5) | 1.08 ± 0.08 | 0.21 ± 0.18 | 1.02 ± 0.08 | 0.04 ± 0.18 |
| **Antibiotics** |  |  |  |  |  |
| Amoxicillin | 77 (4.7) | 1.16 ± 0.11 | 0.40 ± 0.23 | 1.08 ± 0.11 | 0.21 ± 0.23 |
| Piperacillin | 85 (5.2) | 1.37 ± 0.11 | 0.95 ± 0.22 | 1.10 ± 0.13 | 0.25 ± 0.22 |
| Linezolid | 86 (5.2) | 1.56 ± 0.10 | 1.43 ± 0.22 | 1.24 ± 0.12 | 0.60 ± 0.22 |
| Azithromycin | 950 (57.8) | 0.96 ± 0.04 | -0.11 ± 0.10 | 0.94 ± 0.03 | -0.16 ± 0.10 |
| Ceftriaxone | 1024 (62.2) | 1.09 ± 0.04 | 0.22 ± 0.10 | 1.01 ± 0.03 | 0.02 ± 0.10 |
| Meropenem | 139 (8.4) | 1.47 ± 0.08 | 1.20 ± 0.18 | 1.22 ± 0.10 | 0.56 ± 0.17 |
| Levofloxacin | 405 (24.6) | 1.10 ± 0.05 | 0.25 ± 0.11 | 1.04 ± 0.05 | 0.10 ± 0.11 |
| **Immunomodulators** |  |  |  |  |  |
| Hydroxychloroquine | 1498 (91.1) | 0.79 ± 0.08 | -0.54 ± 0.17 | 0.84 ± 0.08 | -0.42 ± 0.17 |
| Tocilizumab | 309 (18.8) | 1.18 ± 0.06 | 0.46 ± 0.12 | 0.97 ± 0.06 | -0.07 ± 0.12 |
| Interferon beta 1-b | 96 (5.8) | 1.22 ± 0.10 | 0.56 ± 0.21 | 1.02 ± 0.12 | 0.04 ± 0.21 |

### Appendix 3: Differences in the characteristic distributions in Table 1 and Table A2a

**Table A3:** The table shows the p-values of testing if the distributions of the patients that underwent end-point events and those who did not are similar.

| Characteristics | *p-values* |  |  |  |  |  |
| --- | --- | --- | --- | --- | --- | --- |
|  | Simple end-point event |  |  | Composite end-point event |  |  |
|  | Whole population | Age (60 - 79) | Age ≥80 | Whole population | Age (60 - 79) | Age ≥80 |
| Initial vital signs - *p-value* |  |  |  |  |  |  |
| Body temperature - °C | $P = .81$ | $P = .07$ | $P = .28$ | $P = .01$ | $P = .01$ | $P = .11$ |
| Heart rate - beats/min | $P = .07$ | $P = .04$ | $P = .27$ | $P = .81$ | $P = .01$ | $P = .15$ |
| Oxygen saturation - % | $P < .001$ | $P < .001$ | $P < .001$ | $P < .001$ | $P < .001$ | $P < .001$ |
| Systolic blood pressure - mm Hg | $P = .79$ | $P = .16$ | $P = .09$ | $P = .80$ | $P = .06$ | $P = .12$ |
| Diastolic blood pressure - mm Hg | $P < .001$ | $P = .08$ | $P = .41$ | $P < .001$ | $P = .04$ | $P = .59$ |
| **Initial laboratory tests - $p$-value** |  |  |  |  |  |  |
| Lactate dehydrogenase - U/L | $P < .001$ | $P < .001$ | $P < .001$ | $P < .001$ | $P < .001$ | $P < .001$ |
| D-dimer - ng/mL | $P < .001$ | $P < .001$ | $P = .02$ | $P < .001$ | $P < .001$ | $P = .04$ |
| Ferritin - ng/mL | $P < .001$ | $P < .001$ | $P < .001$ | $P < .001$ | $P < .001$ | $P = .001$ |
| C-reactive protein - mg/L | $P < .001$ | $P < .001$ | $P < .001$ | $P < .001$ | $P < .001$ | $P < .001$ |
| Lactate - mmol/L | $P < .001$ | $P < .001$ | $P < .001$ | $P < .001$ | $P < .001$ | $P < .001$ |
| Creatinine - mg/dL | $P < .001$ | $P < .001$ | $P < .001$ | $P < .001$ | $P < .001$ | $P < .001$ |
| Lymphocyte count per mm<sup>3</sup> | $P < .001$ | $P < .001$ | $P < .001$ | $P < .001$ | $P < .001$ | $P < .001$ |
| Neutrophil count per mm<sup>3</sup> | $P < .001$ | $P < .001$ | $P < .001$ | $P < .001$ | $P < .001$ | $P < .001$ |
| **Past diagnoses - $p$-value** |  |  |  |  |  |  |
| Current smokers | $P = .98$ | $P = .95$ | $P = .41$ | $P = .92$ | $P = .77$ | $P = .43$ |
| Current and past smokers | $P = .51$ | $P = .20$ | $P = .99$ | $P = .87$ | $P = .18$ | $P = .92$ |
| Diabetes | $P < .001$ | $P = .54$ | $P = .02$ | $P < .001$ | $P = .82$ | $P = .01$ |
| Hypertension | $P = .001$ | $P = .97$ | $P = .56$ | $P = .03$ | $P = .38$ | $P = .44$ |
| Hypercholesterolemia | $P < .001$ | $P = .33$ | $P = .59$ | $P = .005$ | $P = .50$ | $P = .74$ |
| Continued use of anticoagulants | $P < .001$ | $P = .005$ | $P = .59$ | $P < .001$ | $P = .03$ | $P = .72$ |
| Continued use of aspirin | $P < .001$ | $P = .06$ | $P = .03$ | $P < .001$ | $P = .04$ | $P = .04$ |
| Chronic obstructive pulmonary disease | $P < .001$ | $P = .20$ | $P = .91$ | $P < .001$ | $P = .17$ | $P = .69$ |
| Cancer | $P < .001$ | $P = .002$ | $P = .36$ | $P < .001$ | $P < .001$ | $P = .40$ |
| Penicillin allergy | $P = .24$ | $P = .88$ | $P = .40$ | $P = .29$ | $P = .90$ | $P = .44$ |
| Other allergies to medication | $P = .65$ | $P = .29$ | $P = .85$ | $P = .61$ | $P = .47$ | $P = .90$ |

### Appendix 4: Propensity-score models area under curves

**Table A4:**
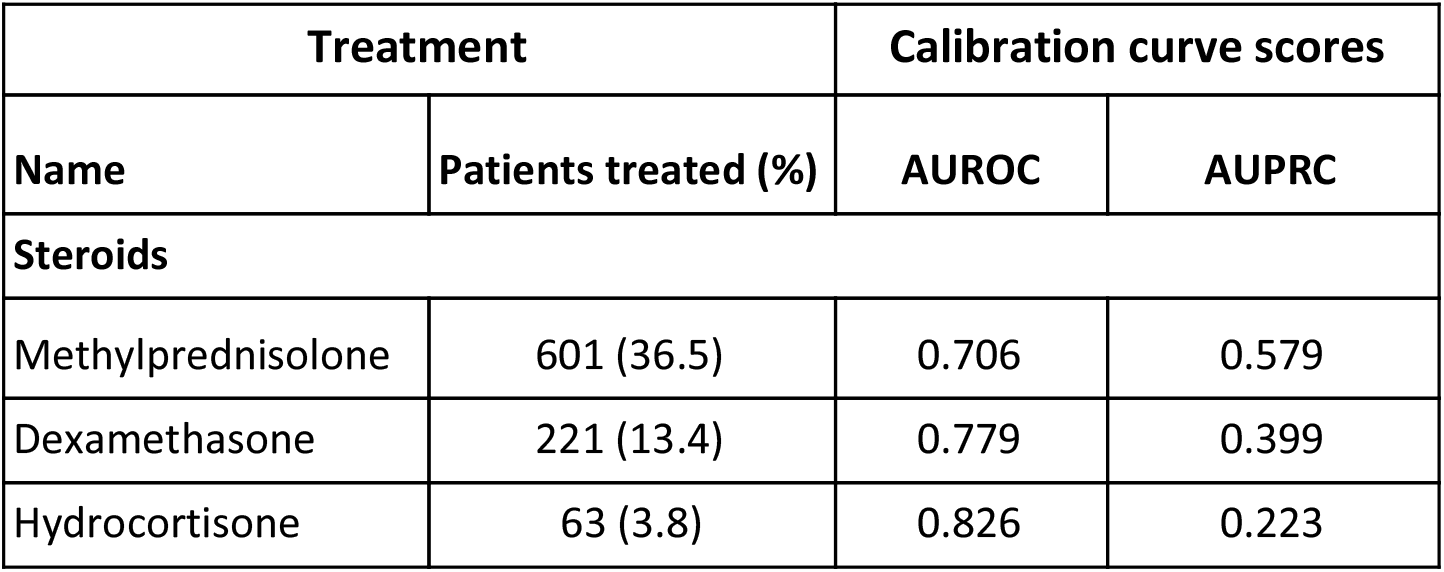
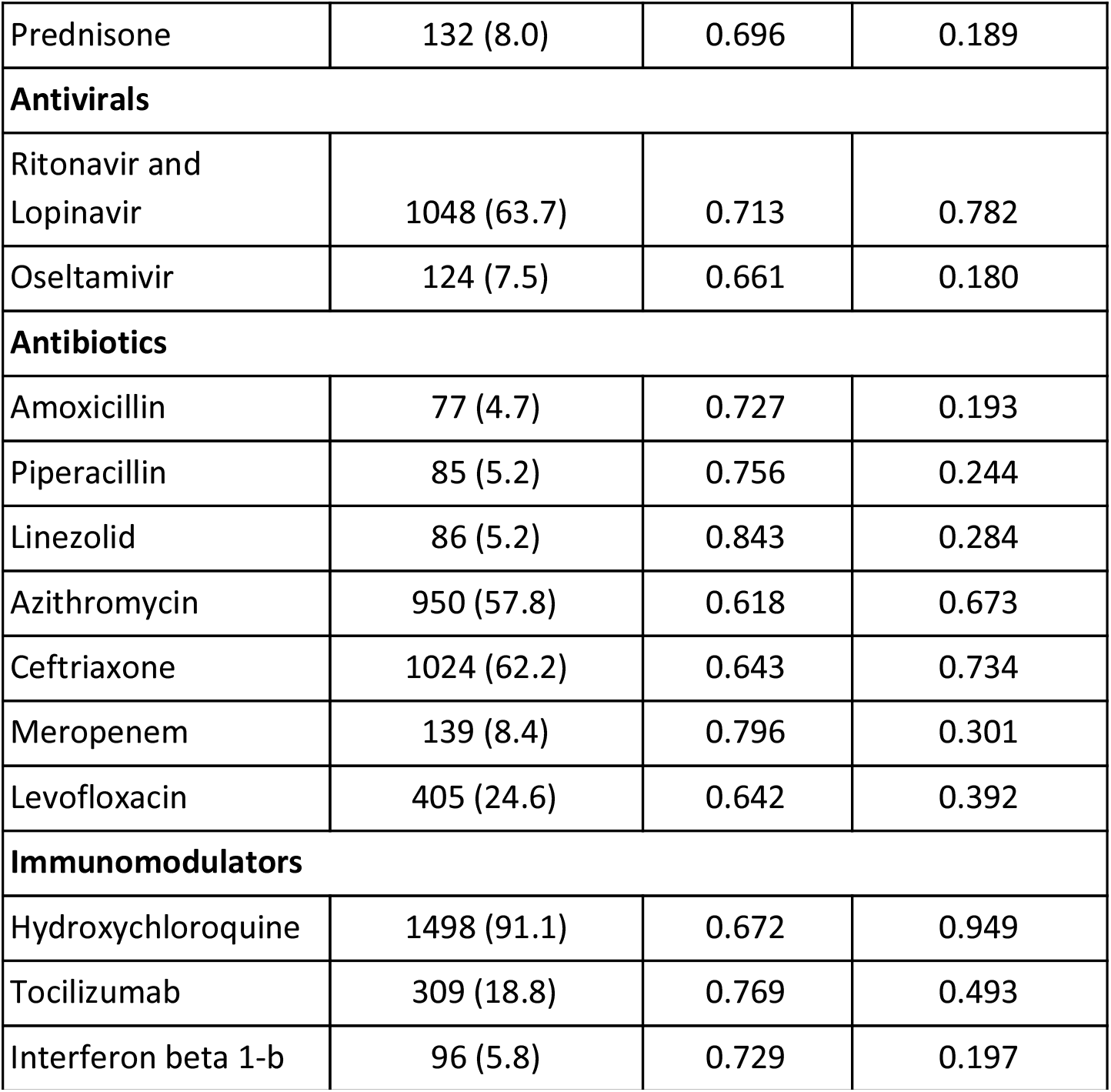
Area under the ROC and Precision-Recall curves for the logistic models for propensity calibration. We present AUPRC in addition to AUROC due to the class imbalance of these classification problems, since it is a better metric here. For a dummy model that predicted the most frequent class with probability equal to its relative frequency, AUROC is 0.5 and AUPRC is the relative frequency of the treated group.

| Treatment |  | Calibration curve scores |  |
| --- | --- | --- | --- |
| Name | Patients treated (%) | AUROC | AUPRC |
| **Steroids** |  |  |  |
| Methylprednisolone | 601 (36.5) | 0.706 | 0.579 |
| Dexamethasone | 221 (13.4) | 0.779 | 0.399 |
| Hydrocortisone | 63 (3.8) | 0.826 | 0.223 |
| Prednisone | 132 (8.0) | 0.696 | 0.189 |
| **Antivirals** |  |  |  |
| Ritonavir and Lopinavir | 1048 (63.7) | 0.713 | 0.782 |
| Oseltamivir | 124 (7.5) | 0.661 | 0.180 |
| **Antibiotics** |  |  |  |
| Amoxicillin | 77 (4.7) | 0.727 | 0.193 |
| Piperacillin | 85 (5.2) | 0.756 | 0.244 |
| Linezolid | 86 (5.2) | 0.843 | 0.284 |
| Azithromycin | 950 (57.8) | 0.618 | 0.673 |
| Ceftriaxone | 1024 (62.2) | 0.643 | 0.734 |
| Meropenem | 139 (8.4) | 0.796 | 0.301 |
| Levofloxacin | 405 (24.6) | 0.642 | 0.392 |
| **Immunomodulators** |  |  |  |
| Hydroxychloroquine | 1498 (91.1) | 0.672 | 0.949 |
| Tocilizumab | 309 (18.8) | 0.769 | 0.493 |
| Interferon beta 1-b | 96 (5.8) | 0.729 | 0.197 |

### Appendix 5: Propensity score distributions

**Figure A5:**
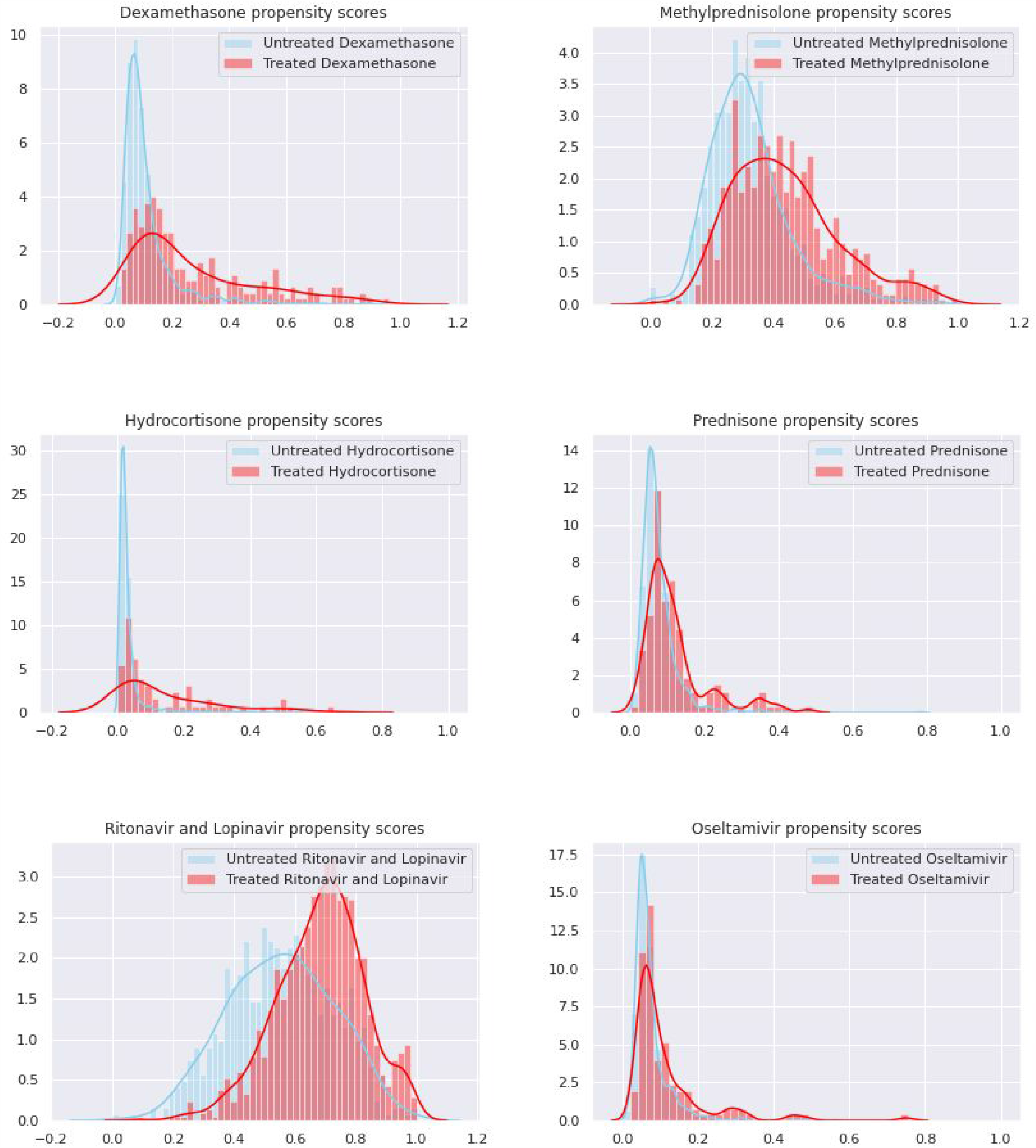
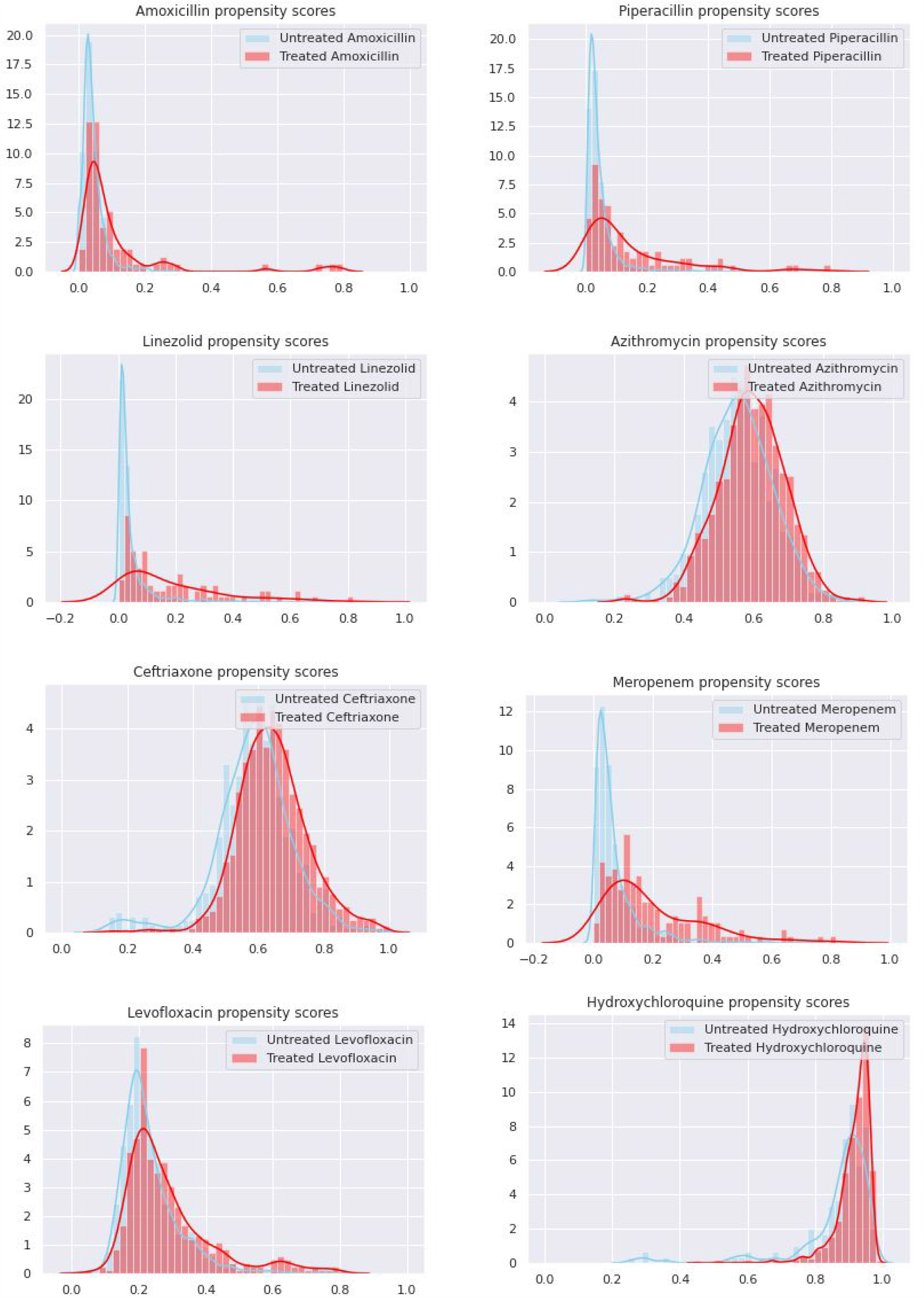
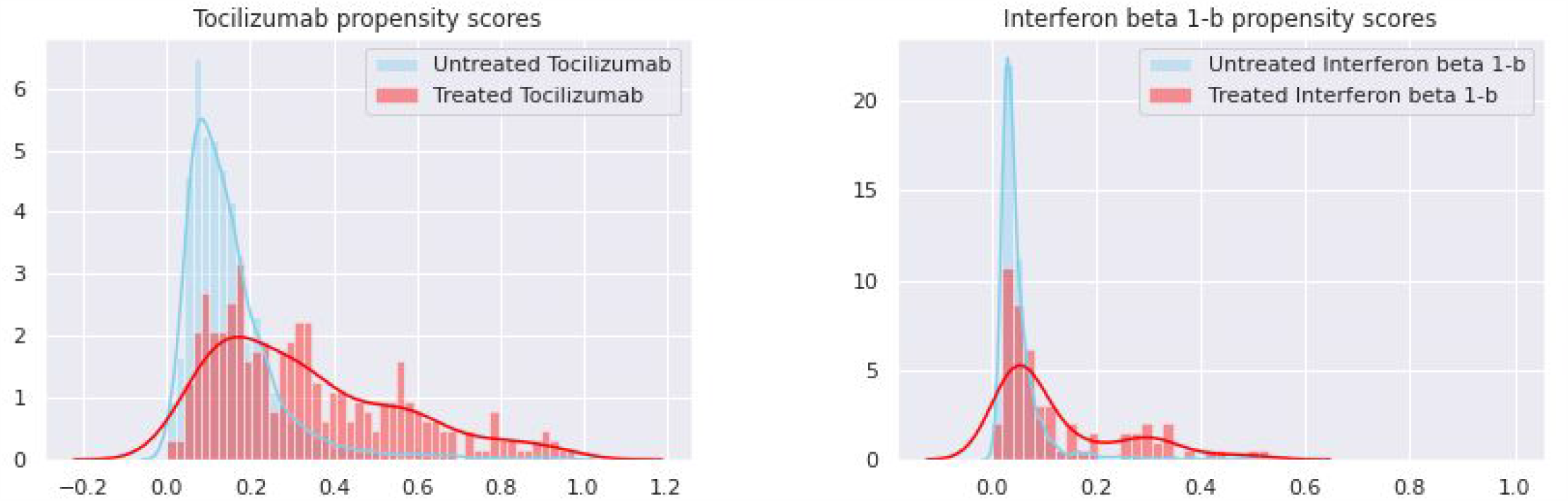
Propensity score distributions for the treated and untreated populations for each of the treatments.

### Appendix 6: Treatment effect with nearest neighbour matching

**Table A6:**
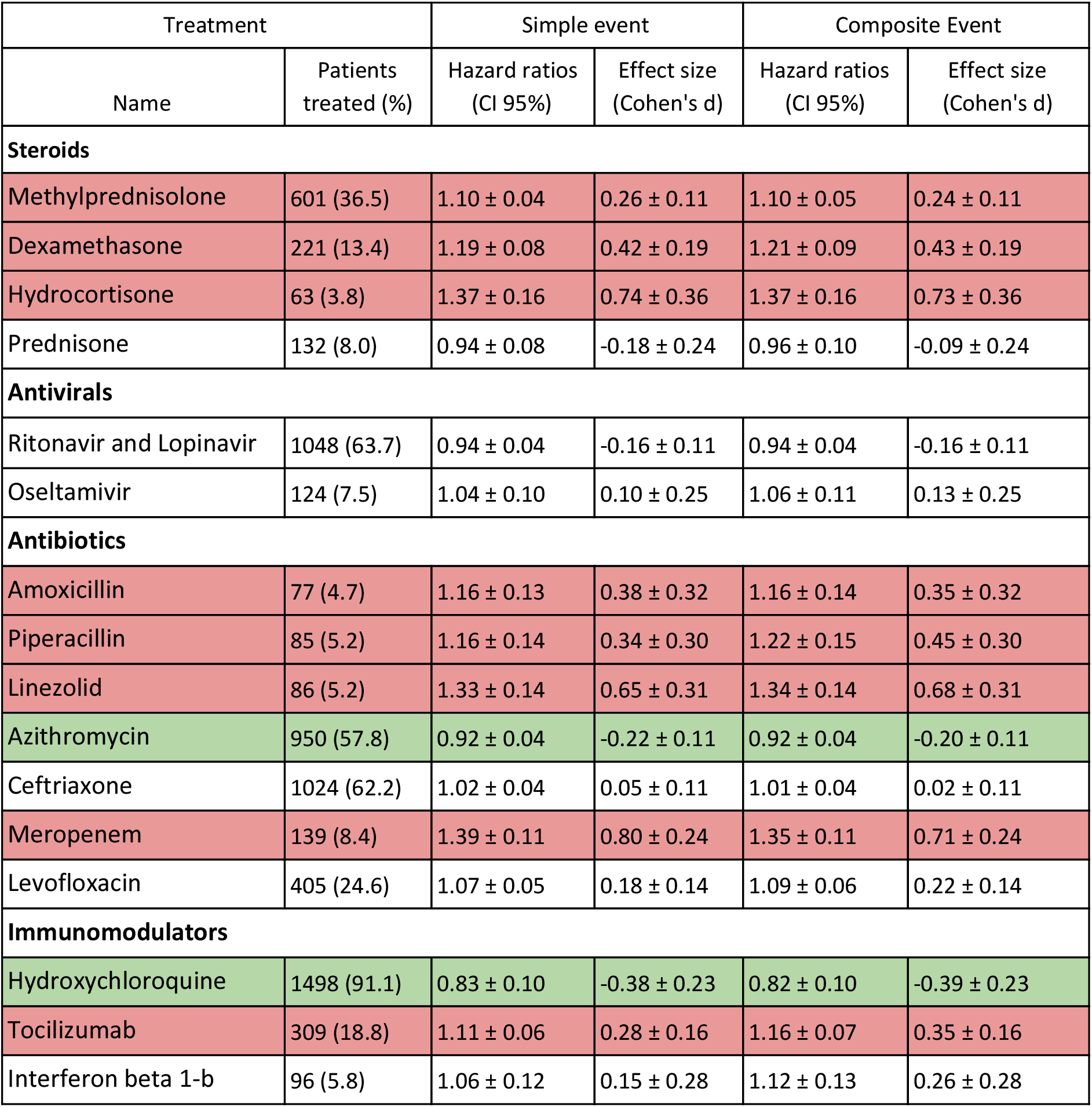
Hazard ratios and effect sizes for NN-matched sample. Every patient in the smaller group was matched to the closest patient in terms of characteristics. (Euclidean distance after normalizing the continuous variables). Medications highlighted in red have at least a small positive d (>0.2) after propensity-score matching. Similarly, medications highlighted in green have d < −0.2.

| Treatment |  | Simple event |  | Composite Event |  |
| --- | --- | --- | --- | --- | --- |
| Name | Patients treated (%) | Hazard ratios (CI 95%) | Effect size (Cohen's d) | Hazard ratios (CI 95%) | Effect size (Cohen's d) |
| **Steroids** |  |  |  |  |  |
| Methylprednisolone | 601 (36.5) | 1.10 ± 0.04 | 0.26 ± 0.11 | 1.10 ± 0.05 | 0.24 ± 0.11 |
| Dexamethasone | 221 (13.4) | 1.19 ± 0.08 | 0.42 ± 0.19 | 1.21 ± 0.09 | 0.43 ± 0.19 |
| Hydrocortisone | 63 (3.8) | 1.37 ± 0.16 | 0.74 ± 0.36 | 1.37 ± 0.16 | 0.73 ± 0.36 |
| Prednisone | 132 (8.0) | 0.94 ± 0.08 | -0.18 ± 0.24 | 0.96 ± 0.10 | -0.09 ± 0.24 |
| **Antivirals** |  |  |  |  |  |
| Ritonavir and Lopinavir | 1048 (63.7) | 0.94 ± 0.04 | -0.16 ± 0.11 | 0.94 ± 0.04 | -0.16 ± 0.11 |
| Oseltamivir | 124 (7.5) | 1.04 ± 0.10 | 0.10 ± 0.25 | 1.06 ± 0.11 | 0.13 ± 0.25 |
| **Antibiotics** |  |  |  |  |  |
| Amoxicillin | 77 (4.7) | 1.16 ± 0.13 | 0.38 ± 0.32 | 1.16 ± 0.14 | 0.35 ± 0.32 |
| Piperacillin | 85 (5.2) | 1.16 ± 0.14 | 0.34 ± 0.30 | 1.22 ± 0.15 | 0.45 ± 0.30 |
| Linezolid | 86 (5.2) | 1.33 ± 0.14 | 0.65 ± 0.31 | 1.34 ± 0.14 | 0.68 ± 0.31 |
| Azithromycin | 950 (57.8) | 0.92 ± 0.04 | -0.22 ± 0.11 | 0.92 ± 0.04 | -0.20 ± 0.11 |
| Ceftriaxone | 1024 (62.2) | 1.02 ± 0.04 | 0.05 ± 0.11 | 1.01 ± 0.04 | 0.02 ± 0.11 |
| Meropenem | 139 (8.4) | 1.39 ± 0.11 | 0.80 ± 0.24 | 1.35 ± 0.11 | 0.71 ± 0.24 |
| Levofloxacin | 405 (24.6) | 1.07 ± 0.05 | 0.18 ± 0.14 | 1.09 ± 0.06 | 0.22 ± 0.14 |
| **Immunomodulators** |  |  |  |  |  |
| Hydroxychloroquine | 1498 (91.1) | 0.83 ± 0.10 | -0.38 ± 0.23 | 0.82 ± 0.10 | -0.39 ± 0.23 |
| Tocilizumab | 309 (18.8) | 1.11 ± 0.06 | 0.28 ± 0.16 | 1.16 ± 0.07 | 0.35 ± 0.16 |
| Interferon beta 1-b | 96 (5.8) | 1.06 ± 0.12 | 0.15 ± 0.28 | 1.12 ± 0.13 | 0.26 ± 0.28 |

### Appendix 7: Caliper size sensitivity and exclusion rates

To ensure that the choice of caliper did not have an outsized effect on the results we repeated the analysis for a range of caliper sizes (from 0.01 to 0.5 as a fraction of the standard deviation). Table A7a shows the median and inter quartile range exclusion rates at each of the caliper sizes tested (we excluded any unmatched patients from the analysis) which justifies using a caliper in the bigger range to avoid excluding too many patients. Table A7b shows the median and interquartile range for the effect size (Cohen’s d) over the caliper range. The results reported in the discussion mostly do not change when taking into account different caliper sizes, except for amoxicillin which at d=0.19 was at the edge of what we considered significant effect and goes to d=0.22 median effect size over the range of calipers.

**Table A7a:** Median exclusion rate for various caliper sizes as a fraction of the standard deviation of the propensity score. The smaller the caliper, the easier it is to have some patients too far away from any other in the opposite treatment group, so the exclusion rate rises. In blue, the caliper size we used for the main analysis.

| Caliper size as fraction of standard deviation | Median exclusion rate (%) [Q1, Q3] |
| --- | --- |
| 0.01 | 17.6 [13.1, 22.6] |
| 0.03 | 10.0 [5.9, 13.1] |
| 0.06 | 6.5 [3.9, 8.9] |
| 0.10 | 3.2 [1.8, 6.4] |
| 0.20 | 1.1 [0.5, 3.2] |
| 0.30 | 0.7 [0.0, 2.1] |
| 0.40 | 0.1 [0.0, 1.8] |
| 0.50 | 0.0 [0.0, 0.9] |

**Table A7b:** Median, Q1 and Q3 effect size over the caliper range for each treatment. Results are very similar to those of Tables 2 and A2b and do not change the conclusions presented. This shows that the results are robust to changing the caliper size.

| Treatment | Median effect over caliper range [Q1, Q3] |
| --- | --- |
| **Steroids** |  |
| Methylprednisolone | 0 [-0.03, 0.01] |
| Dexamethasone | 0.27 [0.24, 0.27] |
| Hydrocortisone | 0.45 [0.44, 0.49] |
| Prednisone | -0.29 [-0.3, -0.28] |
| **Antivirals** |  |
| Ritonavir and Lopinavir | -0.01 [-0.01, -0.05] |
| Oseltamivir | 0.05 [0.04, 0.1] |
| **Antibiotics** |  |
| Amoxicillin | 0.22 [0.21, 0.25] |
| Piperacillin | 0.27 [0.26, 0.29] |
| Linezolid | 0.64 [0.62, 0.72] |
| Azithromycin | -0.16 [-0.17, -0.16] |
| Ceftriaxone | 0.02 [0.01, 0.02] |
| Meropenem | 0.57 [0.56, 0.6] |
| Levofloxacin | 0.097 [0.086, 0.13] |
| **Immunomodulators** |  |
| Hydroxychloroquine | -0.41 [-0.42, -0.4] |
| Tocilizumab | -0.06 [-0.09, -0.03] |
| Interferon beta 1-b | 0.03 [-0.01, 0.06] |

### Appendix 8: Missing data for each variable

**Table A8:** Number of entries in the original dataset for the initial laboratory tests and initial vital signs. Missing data was handled by first using the median to impute and iteratively training multivariate regressions on all the variables except for one to predict the one out. The improved predictions were then used to train new models in the next iteration and this was repeated until convergence. Due to the small sample size, procalcitonin was not used to train the propensity-score models.

| Characteristics | Number of entries (%) |
| --- | --- |
| **Initial vital signs - median [Q1, Q3]** |  |
| Body temperature - °C | 1326 (80.6) |
| Heart rate - beats/min | 1347 (81.9) |
| Oxygen saturation - % | 1359 (82.6) |
| Systolic blood pressure - mm Hg | 1082 (65.8) |
| Diastolic blood pressure - mm Hg | 1082 (65.8) |
| **Initial laboratory tests - median [Q1, Q3]** |  |
| Lactate dehydrogenase - U/L | 1512 (91.9) |
| D-dimer - ng/mL | 1379 (83.8) |
| Ferritin - ng/mL | 486 (29.6) |
| C-reactive protein - mg/L | 1548 (94.1) |
| Lactate - mmol/L | 383 (23.3) |
| Creatinine - mg/dL | 1548 (94.1) |
| Procalcitonin -ng/mL | 102 (6.2) |
| Lymphocyte count per mm<sup>3</sup> | 1569 (95.4) |
| Neutrophil count per mm<sup>3</sup> | 1569 (95.4) |

### Appendix 9: Literature review of treatment effectiveness

Each of the 16 medications in the main analysis were chosen because of their therapeutic relevance during the peak of the pandemic in Madrid. The evidence of their effectiveness was very limited during that time, and is still controversial at most. This is our attempt at collecting the evidence for each of these treatments, both the physiopathologic reasons for them being considered possibly useful in the beginning and a small review of the currently published studies regarding their effectiveness.

#### Steroids

Glucocorticoids were given for their possible role in management of acute respiratory distress syndrome and refractory shock in critically ill patients with COVID-19, apart from their standardised use for exacerbation of chronic obstructive pulmonary disease^1^

During the period when the data was collected (beginning and peak of the pandemic in Madrid), steroids were given very sparingly because of a possibly unfavourable risk/benefit ratio. They were believed to have severe adverse effects in this particular disease, being precisely patients in worse clinical condition those at the highest risk to develop them.

The evidence found in the literature is controversial^2^, but recent results indicate that steroids might have a strong positive effect in patients receiving oxygen or intubated^3^, which would indicate that patients treated with stronger steroids are those with a worse clinical presentation. These results are concordant with the impression of the authors that worked with COVID-19 patients during the pandemic.

#### Antivirals

Ritonavir and lopinavir are viral protease inhibitors used as treatment for HIV. Due to SARS-CoV-2 being a retrovirus that requires similar enzymes to complete its intracellular cycle, they were hypothesized to have some effect in reducing viral load. However, no effect on clinical outcome has been observed in several studies ^4,5^.

Oseltamivir was used with almost no evidence to support it, but it was an antiviral readily available in hospitals. To the best of our knowledge, there is no mechanism through which it could work and studies that have tested its effectiveness have found negative results ^5,6^.

#### Antibiotics

Antibiotics are protocolarily given to patients with viral pneumonia as prophylaxis for bacterial superinfection, a common and grave complication. Amongst them only azithromycin has been hypothesized to have a direct benefit in COVID-19, because of its immunomodulating effect through inhibition of the interleukin-6 inflammatory pathway^7^. There have been some studies that have tested its effectiveness in association with hydroxychloroquine, with discrepant results^8^.

#### Immunomodulators

Hydroxicloroquine was reported to inhibit SARS-CoV-2 in vitro ^9^ and heralded from the beginning of the pandemic as a working treatment for COVID-19 patients. Many studies, mostly observational, have been done to test its effectiveness with very mixed results. However, we agree with Saibal et al.^10^ in that most studies have major methodological limitations, many of them with very small sample sizes and some of the bigger ones with no attempt to control for any confounders ^11^, suggesting that the evidence is very poor. The biggest studies have found no significant effects ^10,12,13^. However the evidence is still unclear. RCTs are ongoing to better assess its effectiveness.

Markedly elevated inflammatory markers (eg, D-dimer, ferritin) and elevated proinflammatory cytokines (including interleukin-6) are associated with critical and fatal COVID-19. Usage of tocilizumab to block the inflammatory pathway has been hypothesized to prevent disease progression^14^. Preliminary studies with small sample sizes seem to suggest positive effects but only in patients at risk of cytokine storm. There are ongoing RCTs to clarify the effects of tocilizumab^15^.

Interferon beta 1-b was suggested to have in vitro activity against Middle East respiratory syndrome coronavirus (MERS-CoV) and good outcomes in an animal model of MERS-CoV infection^16^. Several interferons have been studied in Covid-19 patients. Dastan et al.^17^ found positive results with interferon beta 1a in a non-controlled trial, and interferon beta 1b showed significant improvement in virological and clinical outcomes in a phase 2 trial, both in critical and non-critical patients^18^.

